# Predictors of loss to follow-up among patients receiving antiretroviral therapy in Njombe Region, Tanzania, 2017–2021

**DOI:** 10.64898/2026.02.28.26347333

**Authors:** Hillary Mushi, Meshack D. Lugoba, Raphael Z. Sangeda, Ritah F. Mutagonda, James Mwakyomo, George Musiba, Veryeh Sambu, Beatrice Mutayoba, Mercy Mpatwa, Prosper Njau, Werner Maokola

## Abstract

**Background:** Loss to follow-up (LTFU) undermines the success of antiretroviral therapy (ART) programs, especially in high HIV prevalence regions like Njombe, Tanzania. Understanding factors influencing LTFU is critical to enhance patient retention.

**Aim:** To assess the prevalence and predictors of LTFU among people living with HIV (PLHIV) receiving ART in Njombe, Tanzania, from 2017 to 2021

**Methods:** We conducted a retrospective cohort study using the National Care and Treatment Clinic (CTC2) database, defining LTFU as absence from care for 180 days or more. Logistic regression identified factors associated with LTFU. Data were cleaned using Microsoft Excel and analyzed using IBM SPSS Statistics version 26. Descriptive statistics were used to summarize demographic and clinical characteristics, and logistic regression was used to identify independent predictors of LTFU

**Results:** Of the 37,642 PLHIV initiated on ART, 13,411 (35.6%) were LTFU during the five-year study period. The highest annual incidence of LTFU occurred in 2020 (n = 4,069), coinciding with the onset of the COVID-19 pandemic. District-level disparities were substantial: Wanging’ombe recorded the highest disengagement prevalence (46.7%), while Makete recorded the lowest (23.7%). Multivariable analysis revealed that gender and age were not independent predictors of attrition (p > 0.05). However, significant associations with LTFU were observed for lower pharmacy refill adherence, marital status (single and divorced), and district of residence. Notably, patients in Wanging’ombe had more than double the odds of LTFU compared to those in Njombe (AOR 2.09; 95% CI: 1.95–2.24), whereas the 2021 initiation cohort demonstrated a significantly lower risk of disengagement (AOR 0.25; 95% CI: 0.22–0.28).

**Conclusion:** LTFU remains a critical challenge in the Njombe Region. Targeted interventions, including strengthened pharmacy refill monitoring, district-specific strategies, and psychosocial support for PLHIV, are essential to improve retention and sustain progress toward national HIV treatment goals.

## 1. Introduction

Antiretroviral therapy (ART) has transformed HIV infection into a manageable chronic condition, substantially reducing morbidity and mortality rates worldwide [1–3]. Global ART expansion from 7% coverage in 2005 to over 62% by 2018 has prevented millions of AIDS-related deaths [2]. However, the effectiveness of ART programs depends on continuous engagement in the care process. Loss to follow-up (LTFU), defined in Tanzania as failure to return for ART refill within ≥90 days without documented death or transfer [2], remains a critical barrier to long-term treatment success in many low- and middle-income countries [4–6].

Sub-Saharan Africa, which carries approximately 70% of the global HIV burden [6], experiences some of the highest LTFU rates globally. Reviews have reported that nearly 40% of patients disengage from care within the first years of ART, with retention at two years averaging only 60% [3]. Across the region, LTFU has been linked to adverse clinical outcomes, including viral rebound, CD4 decline, drug resistance, and increased mortality [7–9]. Predictors frequently cited include younger age, male gender, advanced disease, low adherence, socioeconomic vulnerability, and barriers to accessing health facilities [2,4,10–14].

Medication adherence, often assessed using measures such as the medication possession ratio (MPR) or pharmacy refill, is crucial for maintaining viral suppression in HIV patients. Evidence from studies showed a strong relationship between decreasing MPR and increasing risk of virologic failure [9,15], highlighting the importance of early identification of patients at risk of disengagement.

In Tanzania, LTFU continues to undermine ART program outcomes, particularly in regions with high HIV prevalence, including those with significant mining or pastoralist activities [16]. The Njombe Region consistently reports the highest HIV prevalence nationally, making uninterrupted ART engagement essential for controlling transmission and achieving epidemic targets. Despite this epidemiological significance, evidence on district-level patterns of LTFU in Njombe and the demographic and clinical factors associated with disengagement remains limited. While previous studies in Dar es Salaam and other African settings have demonstrated increased risk of LTFU among males, individuals with advanced HIV disease, and those with very low CD4 counts [14], it is unclear whether these findings are applicable to the Njombe context.

Therefore, this study aimed to quantify the magnitude of LTFU, describe its spatial distribution across Njombe’s four districts, and identify demographic and clinical predictors of disengagement from care between 2017 and 2021. The findings are intended to provide region-specific evidence to inform targeted interventions in Tanzania’s highest-prevalence HIV setting.

## 2. Methodology

### 2.1 Study design

This retrospective, facility-based study analyzed routinely collected patient data from the national Care and Treatment Clinic (CTC2) database managed by the National AIDS and Sexually Transmitted Infections Control Programme (NASHCOP) in Tanzania. The CTC2 platform is used nationwide to document HIV care, including demographic characteristics, treatment initiation, ART regimens, pharmacy refills, viral load measurements, and clinical outcomes. For this analysis, all available records for the period 1 January 2017 to 31 December 2021 were extracted and analysed.

### 2.2 Study area

The study was conducted in the Njombe Region, located in the southern highlands of Tanzania, which consistently reports among the highest HIV prevalence rates nationally. Administratively, Njombe comprises four districts: Njombe, Wanging’ombe, Makete, and Ludewa, each served by multiple health facilities that provide HIV care and treatment. These districts vary in terms of population distribution, health service coverage, and geographic accessibility.

### 2.3 Study population

The study population included all individuals enrolled for HIV care and treatment within the Njombe Region during the review period who had at least one documented clinic visit or ART refill in the CTC2 system. Patients lacking essential demographic information or ART initiation dates, or those transferred out of the region, were excluded. Therefore, the dataset represents a census of all ART PLHIV actively recorded in the four districts over five years.

### 2.4 Data collection and management

Patient-level data were extracted from the national CTC2 server by authorized personnel at NASHCOP and provided in the Microsoft Excel format. The dataset included demographic variables (age, gender, and marital status), programmatic details (ART initiation year, treatment regimen dispensed at each visit, and adherence based on pharmacy refill behavior), viral load results, and follow-up status. After initial cleaning in Excel, the data were imported into Power BI and IBM SPSS Statistics version 26 for transformation, recoding, and statistical analysis. Duplicate entries were removed, internal consistency checks were performed, and missing or implausible values were reviewed. All categorical variables were standardized according to the NASHCOP coding conventions to ensure uniformity across facilities and districts.

### 2.5 Variable definitions and operationalization

The primary outcome of this study was the LTFU rate. In accordance with Tanzanian national guidelines, a PLHIV is typically classified as LTFU if they fail to return for care 90 days or more after their last scheduled appointment without documentation of transfer or death [17,18]. However, to ensure a more conservative estimate of long-term disengagement and to align with our final study observation window (June–December 2021), we applied a more stringent criterion: PLHIV were classified as LTFU if they failed to return for care for 180 days or more after their last scheduled visit. For analytical purposes, the follow-up status was dichotomized into LTFU (1) and not LTFU (0)

The independent variables included demographic factors such as age, gender, marital status, and district of residence. Age was analyzed both as a continuous variable and in categorical groups reflecting distinct clinical and developmental stages: children (0–10 years), adolescents (11–18 years), young adults (19–28 years), adults (29–69 years), and older adults (≥70 years). The clinical variables included the year of ART initiation, mean adherence percentage derived from pharmacy refill data, and viral load suppression, defined according to WHO guidance as < 1000 copies/mL for suppressed and ≥ 1000 copies/mL for unsuppressed. The patient’s final program statuses, including active, transferred out, deceased, LTFU, missed appointment, opted out, or not HIV-positive, were also extracted from the CTC2 system.

Data quality control procedures included screening for missing values, logical inconsistencies (such as ART initiation dates occurring after documented follow-up visits), and outlier values. Duplicate patient identifiers were removed, and all categorical variables were recoded to produce mutually exclusive categories that were suitable for analysis. Continuous variables were examined for distributional abnormalities and transformed or categorized where appropriate.

### 2.6 Data analysis

Descriptive statistics were used to summarize the demographic and clinical characteristics of PLHIV across the four districts. Continuous variables, such as age and pharmacy refill rates, were summarized using means and standard deviations, while categorical variables were presented as frequencies and percentages. Age was analyzed both as a continuous variable and in categorical groups reflecting clinical and developmental stages: children (0–10 years), adolescents (11–18 years), young adults (19–28 years), adults (29–69 years), and older adults (≥70 years). Bivariate associations between independent variables and LTFU were evaluated using Pearson’s chi-square test for categorical data and independent-samples t-tests for continuous measures.

Multivariable analysis was performed using binary logistic regression to identify independent predictors of LTFU. Variables with p-value < 0.20 in the bivariate analysis or those judged to be epidemiologically relevant were entered into the adjusted model. Reference categories were selected based on clinical convention and distributional patterns: 2017 for ART initiation year; the pediatric group (0–10 years) for age; Njombe District for residence; and married status. Adjusted odds ratios (aORs) with 95% confidence intervals (CI) were computed to quantify the strength of association. Model fit was assessed using the Omnibus Test of Model Coefficients, Nagelkerke R², Hosmer–Lemeshow test, and the overall classification accuracy. Statistical significance was set at p < 0.05 (two-sided).

### 2.7 Ethical considerations

This study utilized anonymized secondary data from the National Care and Treatment Clinic (CTC2) database. Permission to access and analyze the data was granted by the National AIDS and Sexually Transmitted Infections Control Programme (NASHCOP) and the Ministry of Health, Tanzania. Ethical approval was obtained from the Muhimbili University of Health and Allied Sciences (MUHAS) Research and Ethics Committee (Reference: DA.25/111/298/01B/183; February 2022). The study was conducted in accordance with national ethical guidelines and the Declaration of Helsinki (2013 revision). The ethics committee waived individual informed consent because the study used routinely collected, de-identified data, thereby ensuring patient confidentiality throughout the analysis.

## 3. Results

### 3.1 Geographic distribution of loss to follow-up

LTFU varied substantially across the Njombe Region from 2017 to 2021. Wanging’ombe District recorded the highest proportion of PLHIV LTFU (47.8%), followed by Njombe (33.2%) and Ludewa (32.1%). Makete reported the lowest LTFU rate (23.7%). These spatial differences highlight the marked geographic variation in treatment retention across the region (Figure 1).

**Figure 1:**
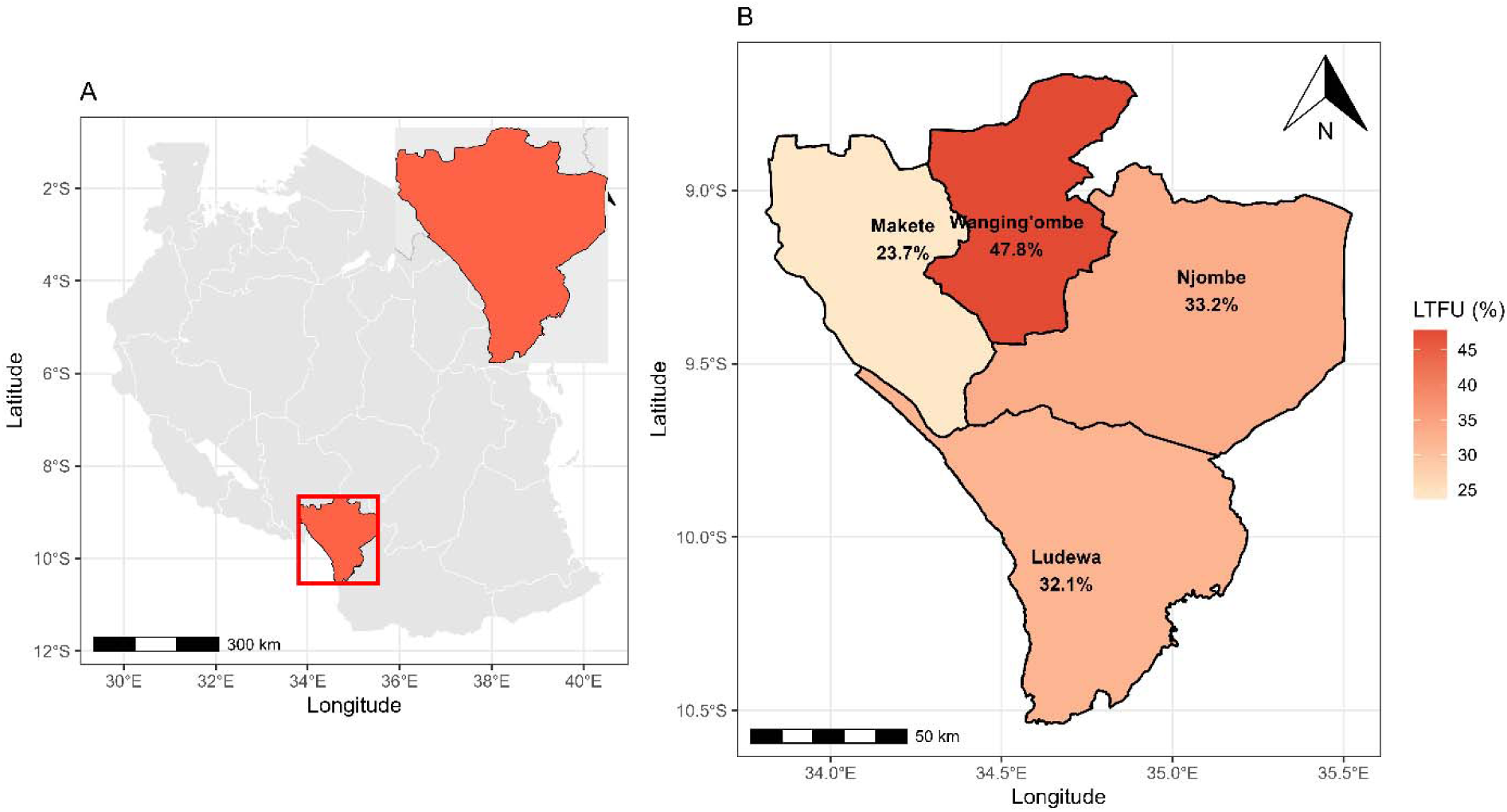
Geographical boundaries of the Njombe Region showing the district-level spatial distribution of loss to follow-up (LTFU) among people living with HIV from 2017 to 2021. Panel A illustrates the location of Njombe in Tanzania, with an inset zooming into the regional boundaries. Panel B presents the district-specific LTFU proportions for Njombe, Wanging’ombe, Makete, and Ludewa districts.

### 3.2 Baseline sociodemographic and clinical characteristics

A total of 39,566 PLHIV from the Njombe, Wanging’ombe, Makete, and Ludewa districts were included in the final dataset. Adults accounted for 93.2% (n = 35,070), while children comprised 6.8% (n **=** 2,572). The mean age of the participants was 36 ± 13 years, and females formed the majority (64.0%, n = 24,087).

Regarding marital status, more than half of the PLHIV were married (57%, n = 21,460). The participants accumulated 888,648 clinic visits between 2017 and 2021. The most frequently dispensed ART regimen was TDF + 3TC + EFV (49.0% of visits), followed by TDF + 3TC + DTG (32.9% of visits).

Njombe District contributed the largest number of PLHIV (54.5%), while Ludewa contributed the smallest (14.5%) (Table 1).

**Table 1:** Baseline demographic and clinical characteristics of PLHIV in Njombe Region, 2017–2021 (N = 39,566)

| Variable | Category | n | % |
| --- | --- | --- | --- |
| **Gender** | Male | 14,418 | 36.4 |
|  | Female | 25,148 | 63.6 |
| **Age group (years)** | 0–10 | 921 | 2.3 |
|  | 11–18 | 1,426 | 3.6 |
|  | 19–28 | 6,317 | 16.0 |
|  | 29–69 | 30,366 | 76.7 |
|  | ≥70 | 536 | 1.4 |
| **Marital status\*** | Married | 23,013 | 58.2 |
|  | Single | 8,038 | 20.3 |
|  | Widowed | 3,068 | 7.8 |
|  | Divorced | 1,374 | 3.5 |
|  | Child | 544 | 1.4 |
|  | Cohabiting | 234 | 0.6 |
| **District of residence** | Njombe | 21,574 | 54.5 |
|  | Wanging'ombe | 6,390 | 16.2 |
|  | Makete | 5,871 | 14.8 |
|  | Ludewa | 5,731 | 14.5 |
| **Year of ART initiation** | 2017 | 22,824 | 57.7 |
|  | 2018 | 4,975 | 12.6 |
|  | 2019 | 5,235 | 13.2 |
|  | 2020 | 3,948 | 10.0 |
|  | 2021 | 2,584 | 6.5 |
| **Viral-load result available** | Yes | 28,218 | 71.3 |
| **Viral suppression (<1000 copies/mL)** | Suppressed | 25,129 | 89.1 |
|  | Unsuppressed | 3,089 | 10.9 |
| **Mean ± SD** | Age (years) | 38.5 ± 13.4 | |
| | Adherence (%) | 86.5 ± 16.3 | |
| | Viral load (copies/mL) | 13 671 ± 239 645 | |

\*Marital-status data are missing for 8.3 % of PLHIV.

### 3.3 Status at last follow-up

At the end of the study period, the clinical status of the cohort was determined based on the most recent recorded visit. Of the total population, 27,011 (68.3%) PLHIV were recorded as remaining active in care. Documented transfers accounted for 16.9% (n=6,674) of the cohort, while 5.0% (n=1,982) were reported deceased. The prevalence of LTFU documents at the last visit was only 5.9% (n=2,334), with an additional 3.6% (n=1,408) of patients having missed scheduled appointments but not yet meeting the formal 90-day criteria for LTFU classification.

### 3.4 Patterns of loss to follow-up across sociodemographic groups

Across the entire cohort, 13,446 PLHIV (34.0%) were lost to follow-up over the five-year period. The LTFU proportions were similar between females (35.7%) and males (35.6%).

Marked yearly variations were observed. LTFU peaked in 2019 (47.3%) and 2020 (46.3%), whereas 2021 showed the lowest proportion (12.5%), reflecting recent improvements in PLWHIV retention.

The district-level variation remained substantial. Njombe District contributed the largest absolute number of LTFU cases (7,010), but the highest proportion was observed in Wanging’ombe (46.7%). Makete consistently had the lowest proportion of children (23.7%). LTFU was more frequent among cohabiting PLHIV compared with other marital categories (Table 2).

**Table 2:** Bivariate associations between selected characteristics and loss to follow-up (LTFU)

| Variable | Category | LTFU n (%) | $\chi^2$ (df) | p-value |
| --- | --- | --- | --- | --- |
|  | **Overall LTFU** | 13,446 (34.0 ) |  |  |
| **Year of ART initiation** |  |  | 1,559.9 (4) | < 0.001 |
|  | 2017 | 29.5 |  |  |
|  | 2018 | 41.9 |  |  |
|  | 2019 | 47.3 |  |  |
|  | 2020 | 46.3 |  |  |
|  | 2021 | 12.5 |  |  |
| **Age group (years)** |  |  | 1,134.0 (4) | < 0.001 |
|  | 0–10 | 33.8 |  |  |
|  | 11–18 | 32.3 |  |  |
|  | 19–28 | 52.3 |  |  |
|  | 29–69 | 30.3 |  |  |
|  | ≥70 | 32.1 |  |  |
| **District** |  |  | 838.5 (3) | < 0.001 |
|  | Njombe | 33.2 |  |  |
|  | Wanging'ombe | 47.8 |  |  |
|  | Makete | 23.7 |  |  |
|  | Ludewa | 32.1 |  |  |
| **Marital status** |  |  | 273.9 (5) | < 0.001 |
|  | Married | 31.7 |  |  |
|  | Single | 40.7 |  |  |
|  | Widowed | 30.0 |  |  |
|  | Divorced | 38.6 |  |  |
|  | Child | 24.6 |  |  |
|  | Cohabiting | 39.7 |  |  |
| **Gender** | Male | 34.0 | 0.03 (1) | 0.87 |
|  | Female | 34.0 |  |  |
| **Viral-load success** | Failure ≥ 1000 | 28.1 LTFU | 209.2 (1) | < 0.001 |
|  | Success < 1000 | 17.4 LTFU |  |  |

### 3.5 Predictors of loss to follow-up

Multivariable logistic regression was conducted to identify factors independently associated with LTFU among PLHIV in the Njombe Region, with the final model demonstrating a strong fit (Omnibus chi-square = 3226.1, p < 0.001; Nagelkerke R^2^ = 0.118). The year of ART initiation emerged as a significant predictor; compared with those who initiated treatment in 2017, patients starting between 2018 and 2020 faced significantly higher odds of disengagement, with the highest risk observed in 2019 (AOR 1.94, 95% CI 1.81–2.07). Notably, initiation in 2021 was strongly protective, associated with a 75% reduction in the odds of LTFU (AOR 0.25, 95% CI 0.22–0.29) relative to the 2017 baseline.

District-level variation remained a robust independent predictor of patient retention. Residents of Wanging’ombe district were more than twice as likely to be lost to follow-up (AOR 2.09, 95% CI 1.96–2.23) compared to those in Njombe. Conversely, PLHIV in Makete had significantly lower odds of LTFU (AOR 0.71, 95% CI 0.66–0.76), resulting in the highest retention levels in the region.

Age and social support structures significantly influenced outcomes. Young adults aged 19–28 years were the most vulnerable demographic, exhibiting more than double the odds of LTFU (AOR 2.07; 95% CI: 1.68–2.55) compared to the elderly reference group (>70 years). Furthermore, marital status was a key determinant of care stability; single (AOR 1.25; 95% CI: 1.18–1.33) and divorced (AOR 1.44; 95% CI: 1.28–1.62) patients had a higher probability of disengagement than their married counterparts (Table 3).

**Table 3:** Bivariate and multivariable logistic-regression analysis of predictors of loss to follow-up among PLHIV in Njombe (N = 36,271)

| Predictor | Univariate<br>COR (95% CI) | p-value | Adjusted OR (95%<br>CI) | p-value |
| --- | --- | --- | --- | --- |
| **Gender** |  |  |  |  |
| Male (Ref) | 1.00 | - | - | - |
| Female | 0.99 (0.95–1.04) | 0.873 | Not Included | - |
| **Year of ART initiation** |  |  |  |  |
| 2017 (Ref) | 1.00 | - | 1.00 | - |
| 2018 | 1.73 (1.62–1.84) | < 0.001 | 1.58 (1.47–1.69) | < 0.001 |
| 2019 | 2.15 (2.02–2.28) | < 0.001 | 1.94 (1.81–2.07) | < 0.001 |
| 2020 | 2.06 (1.92–2.21) | < 0.001 | 1.65 (1.53–1.79) | < 0.001 |
| 2021 | 0.34 (0.30–0.38) | < 0.001 | 0.25 (0.22–0.29) | < 0.001 |
| **Age group** |  |  |  |  |
| 0–10 | 1.00 | - | 1.12 (0.86–1.46) | 0.398 |
| 11–18 | 0.93 (0.78–1.11) | 0.447 | 0.93 (0.73–1.18) | 0.538 |
| 19–28 | 2.15 (1.86–2.49) | < 0.001 | **2.07 (1.68–2.55)** | **< 0.001** |
| 29–69 | 0.85 (0.74–0.98) | 0.024 | 0.86 (0.70–1.05) | 0.135 |
| ≥70 (Ref) | - | - | **Reference** | - |
| **District** |  |  |  |  |
| Njombe (Ref) | 1.00 | - | 1.00 | - |
| Wanging'ombe | 1.85 (1.75–1.96) | < 0.001 | **2.09 (1.96–2.23)** | **< 0.001** |
| Makete | 0.63 (0.59–0.67) | < 0.001 | 0.71 (0.66–0.76) | < 0.001 |
| Ludewa | 0.95 (0.89–1.01) | 0.119 | 0.96 (0.90–1.02) | 0.192 |
| **Marital status** |  |  |  |  |
| Married (Ref) | 1.00 | - | 1.00 | - |
| Single | 1.48 (1.40–1.56) | < 0.001 | 1.25 (1.18–1.33) | < 0.001 |
| Widowed | 0.92 (0.85–1.00) | 0.060 | 1.07 (0.98–1.17) | 0.117 |
| Divorced | 1.36 (1.21–1.52) | < 0.001 | 1.44 (1.28–1.62) | < 0.001 |
| Child | 0.71 (0.58–0.86) | 0.001 | 0.56 (0.44–0.71) | < 0.001 |
| Cohabiting | 1.42 (1.09–1.85) | 0.009 | 1.28 (0.97–1.70) | 0.078 |

**Note:** Omnibus $\chi^2$ (16) = 3226.1, $p < 0.001$ ; Nagelkerke $R^2 = 0.118$ ; Overall correct classification = 68.3%. COR = Crude Odds Ratio; AOR = Adjusted Odds Ratio; CI = Confidence Interval.

### 3.6 Year-by-year burden of LTFU

The annual burden of LTFU fluctuated considerably during the study period. LTFU proportions were highest in 2018–2020, with almost half of the PLHIV lost in 2019, followed by a sharp decline in 2021. Overall, 13,446 PLHIV (34.0%) were lost to follow-up during 2017–2021. The detailed yearly counts and percentages are presented in Table 4.

**Table 4:** Number and percentage of patients lost to follow-up by year, 2017–2021 (N = 39,566)

| Year | Lost to follow-up (n) | % | Not lost (n) | % | Total (n) | % |
| --- | --- | --- | --- | --- | --- | --- |
| 2017 | 6,733 | 29.5 | 16,091 | 70.5 | 22,824 | 100.0 |
| 2018 | 2,086 | 41.9 | 2,889 | 58.1 | 4,975 | 100.0 |
| 2019 | 2,477 | 47.3 | 2,758 | 52.7 | 5,235 | 100.0 |
| 2020 | 1,828 | 46.3 | 2,120 | 53.7 | 3,948 | 100.0 |
| 2021 | 322 | 12.5 | 2,262 | 87.5 | 2,584 | 100.0 |
| **Total** | **13,446** | **34.0** | **26,120** | **66.0** | **39,566** | **100.0** |

## 4. Discussion

With the rapid expansion of ART coverage globally, LTFU remains one of the most significant challenges to the long-term success of HIV/AIDS treatment programs. Patients who discontinue ART are at high risk of rapid viral rebound, CD4 depletion and opportunistic infections, which may lead to early mortality [1,2].

This study assessed the magnitude and predictors of LTFU among PLHIV in the Njombe Region, which has the highest HIV prevalence in Tanzania. Sustaining patient retention during long-term therapy is vital for achieving optimal outcomes. Between 2017 and 2021, 13,411 (35.6%) PLHIV were LTFU, a high proportion that threatens progress toward the UNAIDS 95-95-95 targets. Among these, 1,710 (12.8%) were recorded as deaths, leaving 11,701 (87.2%) as non-death LTFU cases. Our findings showed a difference between the point prevalence of LTFU recorded in the routine CTC2 dashboard (5.9%) and the cumulative incidence calculated over the five-year study period (35.6%). This highlights the complementary value of different reporting metrics. While the national system provides essential real-time status for clinical management, longitudinal cohort analyses offer an additional perspective on long-term retention trends. Such insights are valuable for the NACP in identifying windows for early intervention.

The highest number of LTFU cases occurred in 2020 (n = 4,069), likely reflecting the impact of the COVID-19 pandemic, which caused widespread service disruptions and mobility restrictions [19]. This peak is consistent with trends reported across sub-Saharan Africa, where facility attendance and clinical monitoring were sharply constrained by pandemic-related lockdowns [3]. However, the study also documented a dramatic, paradoxical shift in retention following this peak. While the risk of LTFU was highest during 2019–2020, disengagement declined significantly in 2021. This sharp improvement likely reflects the rapid scale-up of Multi-Month Dispensing (MMD) and other differentiated service delivery (DSD) models implemented by NASHCOP to mitigate pandemic-related disruptions [20]. By reducing the required frequency of clinic visits, these programmatic adaptations likely addressed long-standing structural barriers to retention, such as high transport costs and long facility wait times, thereby stabilizing care for a significant portion of the cohort.

Interestingly, our multivariable analysis found no significant difference in LTFU between males and females (p = 0.852). While earlier literature frequently identified male gender as a risk factor for attrition in certain African cohorts [4], our findings align with more recent regional studies suggesting that gender may not universally predict disengagement [2,14]. This indicates that in the Njombe Region, attrition is likely driven more by structural or geographic factors than by gender-specific behaviors, emphasizing the importance of context-specific retention strategies.

In contrast to gender, age and marital status emerged as critical predictors of retention. Young adults (19–28 years) were significantly more likely to be lost to follow-up than all other cohorts, including both older people and children. This finding is echoed in Ugandan studies, which attribute superior pediatric retention to intensive guardian involvement and reduced stigma among younger children [2]. In Njombe, high attrition was observed among young adults, who faced more than double the odds of disengagement compared to elders. This suggests a transition gap, where the move from pediatric-supported care to independent adult care increases the risk of LTFU [2]. Similarly, single and divorced PLHIV exhibited higher attrition than their married counterparts, likely due to weaker social support networks and more unstable living arrangements. These results align with broader evidence from sub-Saharan Africa identifying adulthood, marital status, advanced disease stage, and tuberculosis (TB) co-infection as primary drivers of LTFU [6,7,15,17]. In many cases, advanced disease contributes to unrecorded deaths within the first three months of the last clinical visit [12]. Furthermore, poor pharmacy refill patterns among patients with advanced immunosuppression may result in a limited treatment response, eventually leading to clinical disengagement [10].

Marked geographic disparities were observed across the study area, with the district of residence serving as a robust independent predictor of retention. Wanging’ombe reported the highest likelihood of LTFU, with more than double the odds of disengagement compared to the regional average, whereas Makete demonstrated the highest retention levels (AOR 0.71, 95% CI 0.66–0.76). These disparities likely reflect variations in service accessibility, population mobility, facility workload, or localized patient-tracing capacity, all of which are recognized drivers of retention in resource-limited settings. The success observed in Makete suggests that factors specific to this district may contribute to higher retention and warrant further investigation for potential adaptation elsewhere. Consequently, shifting from a “one-size-fits-all” approach to district-tailored interventions and localized resource allocation is essential for improving regional ART performance. The drivers of disengagement from HIV care appeared to differ by ecological setting.

In pastoral districts such as Manyara, mobility associated with seasonal livestock movement likely produces episodic facility attendance and apparent LTFU despite continued treatment use [16,21]. In contrast, Njombe represents a long-standing, high-prevalence epidemic where a large proportion of patients have been on antiretroviral therapy for many years. In such mature treatment programs, disengagement could be influenced by factors such as treatment fatigue and normalization of HIV as a chronic condition rather than geographic inaccessibility [22–24]. Previous studies in sub-Saharan Africa have shown that long treatment duration and perceived wellness are important predictors of intermittent engagement in care, particularly in high-coverage settings [22,25].

Unlike studies from Nigeria, Namibia, and other regions of Tanzania that identified male gender as a primary predictor of attrition [4,14], gender was not associated with LTFU in this cohort (p = 0.852). This finding aligns with contemporary East African research, which reports that the effects of gender on retention are highly context-dependent [2]. Such variations underscore the necessity of localized assessments rather than relying on broad regional assumptions; in the Njombe context, structural and geographic factors appear to outweigh gender-specific drivers of disengagement. These insights are vital for designing retention strategies tailored to the specific needs of these patient populations.

Overall, these findings illustrate the multifactorial nature of LTFU in the Njombe Region, shaped by a complex interplay of demographic factors, social circumstances, district-level program characteristics, and system-wide disruptions such as the COVID-19 pandemic [19]. Strengthening patient retention may benefit from targeted, context-specific strategies rather than a generalized approach. Key recommendations include expanding DSD models, enhancing digital patient-tracking systems through NASHCOP, and developing peer-led support mechanisms tailored for young adults [20], enhancing digital patient-tracking systems through NASHCOP, and developing peer-led support mechanisms tailored for young adults, and modelling and usingnhanced language models [26,27]. Furthermore, prioritizing resource allocation and focused interventions in high-burden districts, such as Wanging’ombe, is essential for sustaining the long-term success of the ART program and achieving regional HIV epidemic control.

### Study Limitations and Policy Implications

This study has several limitations that should be considered. First, as a retrospective analysis of routinely collected NASHCOP CTC2 data, the findings were constrained by missing or incomplete records for certain variables. Second, the dataset could not verify silent transfers of patients who may have re-engaged in care at facilities outside the Njombe Region, which potentially leads to an overestimation of true LTFU. Third, because our definition of LTFU required a 180-day window of absence, the lower risk observed in the final study year (2021) likely reflects shorter follow-up duration (right-censoring) compared to earlier cohorts, as these patients had less time to meet the disengagement threshold. Finally, some clinical parameters, such as CD4 count, WHO clinical stage, and tuberculosis co-infection, were not consistently available for all records and could not be assessed as independent predictors.

Despite these limitations, this study provides essential programmatic evidence to guide improvements in ART retention in Tanzania. The significant geographic and demographic disparities observed indicate that retention interventions should be district-specific and socially adaptive. Scaling up DSD models, such as community ART refill groups, and strengthening digital patient-tracking through NASHCOP are vital steps. Furthermore, enhancing psychosocial and peer support services, particularly for young adults and unmarried PLHIV, is essential for improving long-term stability. Implementing such measures may be important for sustaining progress toward national HIV goals and achieving global targets for epidemic control.

## 5. Conclusion

LTFU remains a major barrier to achieving sustained viral suppression and eliminating HIV as a public health threat in Tanzania. Disengagement from care increases the risk of antiretroviral drug resistance, virologic failure, and mortality, thereby undermining progress toward the UNAIDS 95-95-95 targets. In this study, while gender was not an independent predictor of LTFU, marital status, the young adult age and district of residence were significant determinants of retention.

The marked geographic disparities, particularly the high risk of disengagement in Wanging’ombe District, underscore the need to shift from generalized interventions to geographically tailored retention strategies. Strengthening community-based ART delivery models, improving digital client-tracking systems within NASHCOP, and expanding psychosocial and peer support mechanisms for unmarried and young individuals are essential steps to mitigate attrition. Addressing these structural and social drivers is critical for sustaining the long-term success of the ART program in the Njombe Region and providing a framework for similar high-burden, resource-limited settings.

## Declarations

None

## Author Contributions

HM contributed to the study design, data curation, statistical analysis, and drafting of the initial manuscript. RZS conceived the study, provided overall supervision, interpreted the findings, prepared the visualizations, including geospatial mapping, and critically reviewed and revised the manuscript. MDL and JM contributed to the study design and provided methodological inputs. RFM contributed to the clinical interpretation and contextualization of the findings. GM contributed to facility-level data interpretation and clinical programme insights. VS, BM, PN, and WM facilitated data access, programme oversight, and interpretation within the national HIV programme context. MM contributed to policy interpretation and linkage to national disease control strategies. All the authors have reviewed and approved the final manuscript.

## Acknowledgements

The authors thank the National AIDS and Sexually Transmitted Infections Control Programme (NASHCOP) and the Ministry of Health, Tanzania, for granting access to CTC2 data. We also acknowledge the healthcare workers at ART clinics in the Njombe Region for their role in routine data collection.

## Ethics Approval

This study used anonymized secondary data from the National Care and Treatment Clinic (CTC2) database. Ethical approval was obtained from the Muhimbili University of Health and Allied Sciences (MUHAS) Research and Ethics Committee (reference number DA.25/111/298/01B/183; February 2022). Permission to access and analyze the data was granted by the National AIDS and Sexually Transmitted Infection Control Programme (NASHCOP) of the Ministry of Health, Tanzania. Individual informed consent was waived because the study used routinely collected, de-identified data from the hospital database.

## Data Availability Statement

The data supporting the findings of this study are available from the corresponding author upon reasonable request. The data are not publicly available due to ethical restrictions and patient confidentiality.

